# A Randomized Clinical Trial Comparing Visual Inspection with Acetic Acid (VIA) to Pocket Colposcopy for the Triage of HPV+ women living with HIV in Kisumu, Kenya

**DOI:** 10.1101/2024.11.05.24316753

**Authors:** Mary E. Dotson, Eliza Steinberg, Maria Olivia Santos, Jeniffer Ambaka, Megan Huchko, Nimmi Ramanujam

## Abstract

**Objective:** The World Health Organization recommends a “screen, triage, treat” approach for cervical cancer screening for Women Living with HIV (WLWH) in resource-limited settings, with Human Papillomavirus (HPV) testing preferred for screening. We assessed the use of the Pocket colposcope as an adjunct tool to Visual Assessment with Acetic Acid (VIA) for the triage of HPV+ WLWH.

**Methods:** We carried out a randomized clinical trial across six clinics in Kisumu, Kenya between November 2022 and April 2023 (NCT04998318). WLWH who screened positive with self-collected HPV were randomized to either the VIA or Pocket arm. Exam positivity was determined by presence or absence of aceto-white epithelium (AWE). Directed biopsies were performed on AWE; if negative, two random biopsies were taken. Pathology was used to determine diagnostic accuracy. Providers and participants took brief surveys after each exam.

**Findings:** The rate of a positive exams was 17.3% for VIA compared to 14.3% for the Pocket. The overall rate of CIN2/3 was 15.4%, with 12.2% in the VIA Arm and is 18.8% in the Pocket Arm. Pocket and VIA performed comparably on all sensitivity, specificity and negative predictive value (NPV). For Pocket compared to VIA, Sensitivity was 26.3% vs 25.0%; specificity was 88.9% vs 84.0%; and NPV was 82.9% vs 87.1%. However, the positive predictive value (PPV) of the Pocket colposcope arm was almost a factor of two higher than that of the VIA arm (Pocket arm PPV was 375 is and that of the VIA arm was 20.6%). The Pocket Colposcope was acceptable to providers and patients for clinic-based triage of HPV positivity.

**Conclusion:** Provider assessment with the Pocket colposcope detected significantly more treatable disease, thereby reducing the need for overtreatment. This study indicates that the Pocket colposcope is a feasible, lower cost colposcopic device, which could facilitate biopsy-confirmation of disease, increase provider training, patient education and facilitate remote diagnosis.

## Introduction

Cervical cancer remains a global health challenge because effective prevention technologies are not accessible to most women around the world. Each year, almost 600,000 women worldwide are diagnosed with cervical cancer, and over 330,000 women die from the disease.(1) Invasive cervical cancer is highly preventable through vaccination for the human papillomavirus (HPV) or screening, diagnosis, and treatment of cervical precursor lesions.(2) The highly effective prevention methods that are the standard in most developed countries are not widely available in low- and middle-income countries (LMICs) due to cost and infrastructure requirements. To avert deaths of up to 50 million women who will not have effective coverage from the vaccine in the next fifty years, the World Health Organization (WHO) recommends adoption of simplified screening technologies coupled directly with treatment.(3) One such strategy, molecular testing for HPV, has been shown to reduce the incidence and mortality from cervical cancer when coupled directly with outpatient ablative treatment for women with HPV-positive results.(4) However, this “screen & treat” approach leads to overtreatment with consequent burdens on both women and health care systems, and potential under-treatment of missed cancers.(5) These risks are increased among women living with HIV (WLWH), and as a result, the WHO recently updated their recommendations to include a triage step for this population.(6) Kenya, a middle-income country with a high burden of cervical cancer and HIV, has also adopted guidelines that recommend HPV with a triage step prior to treatment.(7)

The currently recommended triage tests, colposcopy or visual inspection with acetic acid (VIA), are not effective solutions for HPV positive women in LMICs. Colposcopy with biopsy is the standard of care for positive screening tests in most high-income settings. Colposcopes are expensive and require a second visit to a referral facility, a trained provider, a pathologist, and a concurrent tracking system for patients, specimens and results. All these factors make colposcopy and biopsy an unrealistic triage method for many women in LMICs. VIA, a simpler and less expensive test, is often used instead. VIA involves applying acetic acid to the cervix, examining the cervix with the naked eye, followed by a diagnosis and treatment decision. VIA has high variability depending on provider experience, patient age, and size of lesion and has low sensitivity due to lack of adequate lighting and magnification.^2-7^ Because there is no ability to capture images with VIA, there is no mechanism for quality monitoring or opportunity for skills improvement.(8)^-^(9) There remains a crucial need for a triage technique that is low-cost, easy-to-use and will accurately identify precancerous lesions.

To address these gaps, our team developed a low-cost, mobile colposcope, called a Pocket Colposcope.(10)^-^(11) The device is powered by a cell phone which houses a software called the Calla Health App (CHA) through which patient information and corresponding images can be captured, stored, and transferred between health facilities or to remote specialists for diagnostic evaluation. The Pocket is inserted through a speculum and positioned at a short working distance (3 cm) to the cervix, obviating the need for high-end optics and high-resolution cameras used in long working distance (30 cm) standard Colposcopes. The Pocket Colposcope has image quality comparable with standard colposcopes and has the capability to perform both white light imaging of acetic acid and green light imaging.(12)

The Pocket colposcope has been tested in more than 6 countries (most of which are LMICs), and has been successfully used by physicians, nurses, and clinical officers. The Pocket colposcope can be used to bridge the gaps in health care infrastructure and services. Specifically, it can be used by primary providers as an adjunct to improve visual triage for women who are HPV positive or can be coupled with biopsy and used as a lower cost replacement to colposcopy. The CHA software can provide linkages between providers and care facilities, facilitating remote diagnosis and reducing attrition to completion of care. Further, the images can be stored in a database that can be retrieved for training of deep learning algorithms which can be used to facilitate diagnosis either as a standalone tool or as an aid to a physical provider.

We sought to evaluate the effectiveness of the Pocket as an adjunct to visual inspection through a randomized comparison of diagnostic accuracy of standard VIA to that of the Pocket colposcope for the triage of HPV positive WLWH in Western Kenya (Figure 1). To evaluate implementation readiness, we compared provider and patient experiences with the VIA and Pocket. **Figure 1**. Pocket Colposcope and Calla Health App Integration.

**Figure 1.**
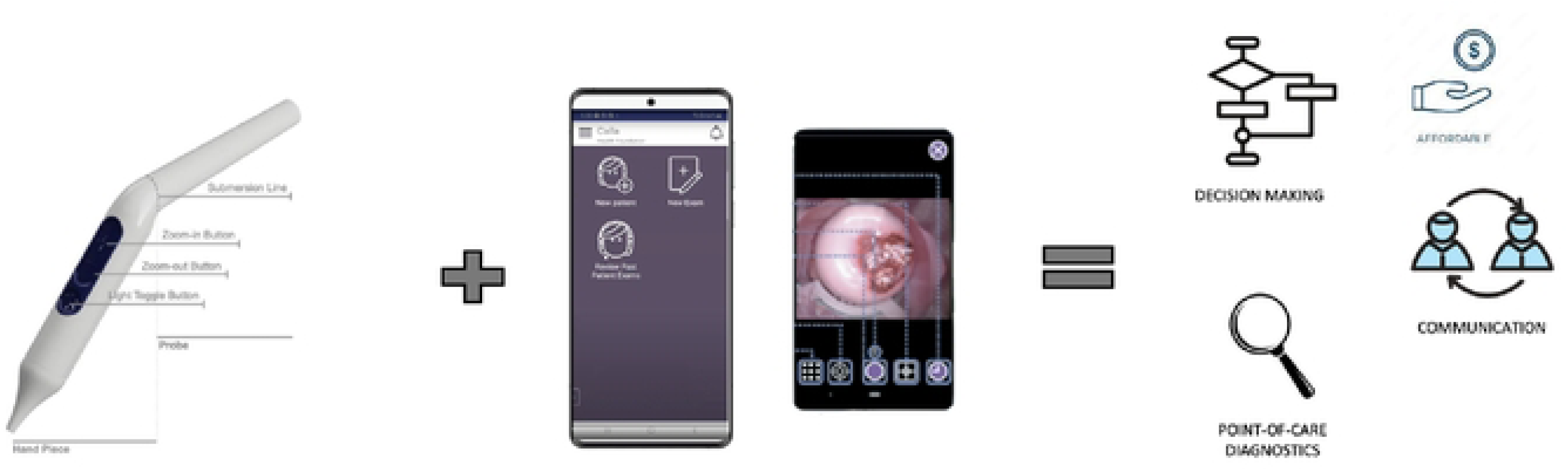

## Methods

### Study Site

This study took place in 6 clinics providing HIV care in the Municipality of Kisumu, Kenya from November 2022 to June 2023. Kisumu is in the Nyanza Province in western Kenya, consisting of a population approximately 500,000 people. It contains both rural and urban households.

### Study Design

We carried out an RCT to examine the diagnostic accuracy of the Pocket Colposcope as a triage tool for cervical cancer screening. WLWH were approached across six clinics to participate in HPV testing using self-collected cervical sampling. Upon receiving a positive HPV test result, women were recruited to the study if they were between 25 and 65 years of age and planned to undergo evaluation for treatment at the clinic following their positive HPV test result. Women were excluded if they declined to consent to the study, were pregnant, had a history of cervical cancer, or had a pelvic exam with concern for infection or invasive cervical cancer.

Women who consented to the study were randomized to either the standard-of-care (VIA) arm or the intervention (Pocket Colposcope) arm. For women in both study arms, providers applied acetic acid to the cervix and waited 60 seconds before visualizing the cervix for any aceto-whitened lesions. For women randomized to the standard-of-care arm, providers performed standard VIA. In the Pocket arm, providers used the Pocket Colposcope to visualize the cervix before and after acetic acid application and to capture images using both white and green light (Figure 2). **Figure 2**. Enrollment of 404 HPV+ women living with HIV, were randomized to receive the control arm (VIA) or the intervention arm (enhanced VIA using the Pocket Colposcope). An imbalance in participant numbers in each arm occurred due to a technical error in the randomization scheme.

**Figure 2.**
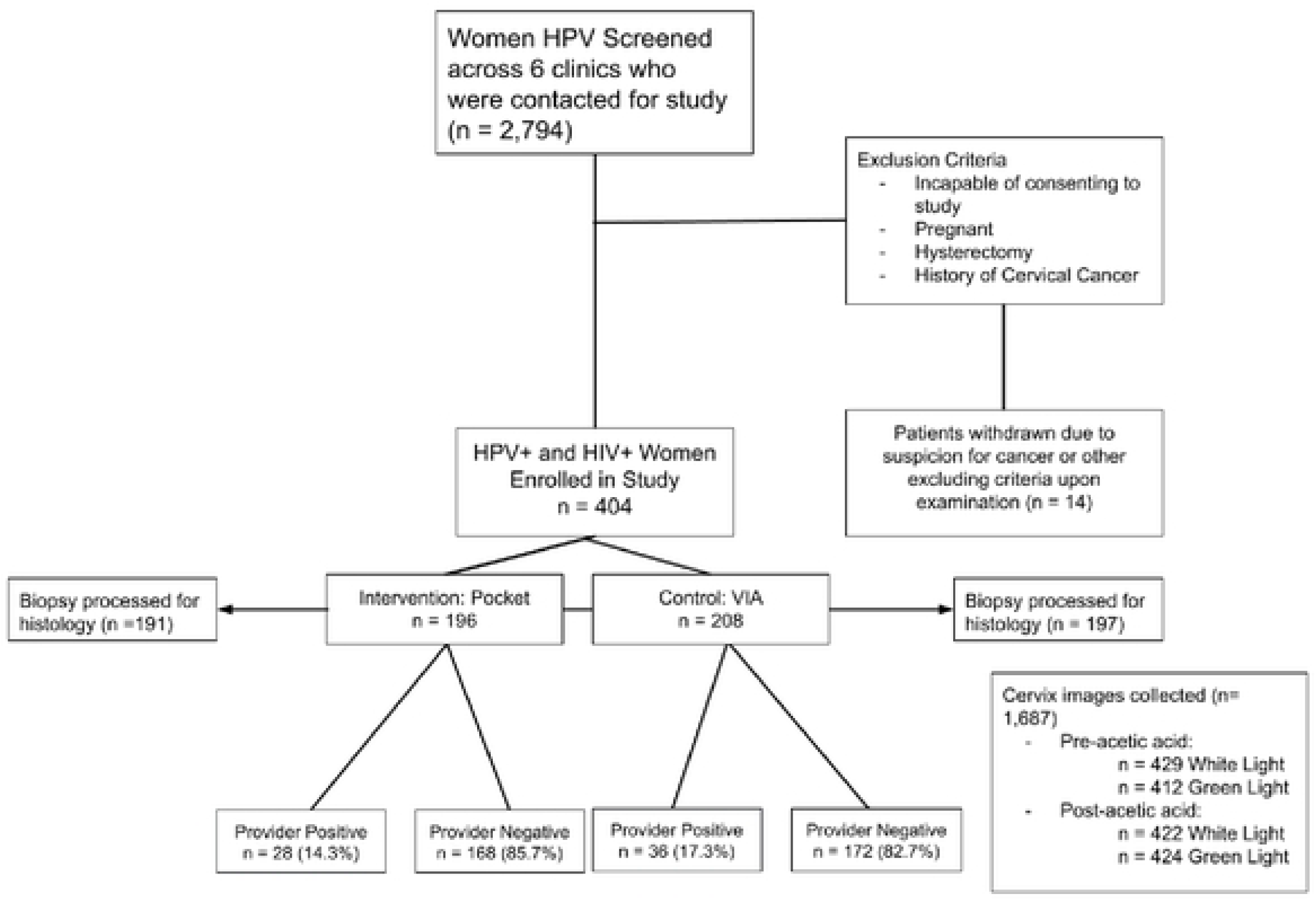

In both arms, providers provided a diagnosis of either “positive” if they believed there was an area of aceto-white epithelium (AWE) present after a minute or “negative” if no changes were seen. Women in both arms had biopsies taken in areas with AWE, or if no AWE was visualized, providers took biopsies at two random locations. Per WHO recommendations all HPV screen-positive women eligible for ablative treatment received treatment in the clinic, regardless of the provider’s visual diagnosis.(7) Participants who were suspected to have lesions too large to treat (using VIA or Pocket), or areas with suspected invasive carcinoma were referred for treatment at an advanced care center. They were withdrawn from enrollment.

Following the exam, a member of the research team administered a questionnaire to assess the participant’s experiences with VIA or the Pocket Colposcope, knowledge of cervical cancer before and after the exam, comfort, and pain during the exam, perceived confidence in the provider’s diagnosis, and the likelihood of recommending the exam to a friend. Providers filled out surveys after each exam to assess their experiences with VIA or the Pocket Colposcope as it related to their ability to complete the exam, ability to make a diagnosis, perception of patient comfort, experience with patients sharing any discomfort, concerns, or pain during the exam, with opportunity for qualitative elaboration, and experience with any challenges during the exam.

### Provider and Study Staff Training

Kenyan providers were trained on using the Pocket Colposcope and CHA using the “train the trainer” method. A clinical officer with 5 years of training and experience performing VIA partnered with staff from Duke University to provide clinical and device training to the KEMRI research assistants, nurses, and community health volunteers prior to the start of enrollment. The training covered basic HPV and Cervical cancer background best practices for VIA, the Pocket Colposcope (capturing high-quality images with the Pocket Colposcope) and CHA in clinical exams and optimizing clinic workflow. Following the in-person training, KEMRI study staff trained local providers on operating the Pocket Colposcope and the accompanying CHA app. Providers were clinical officers and nurses providing general outpatient and HIV-specific care in government of Kenya clinics. Study providers incorporated study visits into their regular clinical duties. Duke study staff maintained weekly calls with KEMRI research assistants throughout the course of the study to troubleshoot any issues.

### Data Management and Analysis

All exam forms, participant and provider surveys, and cervix images were documented and saved in CHA on a mobile device and were later uploaded to a HIPAA-compliant online database. Histopathological results from cervical biopsies were later matched to each corresponding participant identification number. Data were analyzed using R (version 2022.07.02) to determine the concordance between provider diagnoses and histopathology. Histopathological results were used as the “gold standard” when calculating, sensitivity, specificity, positive predictive value (PPV), and negative predictive value (PPV) for provider diagnoses in each study arm. For participant and provider surveys, frequencies and percentages are presented for each answer choice.

### Ethical Review

The study protocols, consent forms, and provider and patient questionnaires were reviewed by the institutional review boards of both the Kenya Medical Research Institute (KEMRI) and Duke University prior to study initiation.

## Results

### Demographics

Between November 2022 and April 2023, 2,794 women living with HIV screened for HPV, 503 (18%) tested HPV-positive, and 404 HPV-positive women were enrolled, randomized, and underwent triage with VIA (208) or the Pocket colposcope (196), which will be referred to the Pocket from this point on (Figure 2). The median age of the participants was 39 years old (IQR: 28-65). The largest number of patients were enrolled at Lumumba, a sub-county hospital with the greatest patient volume among the six clinics (Table 1). Majority of women had three or more children, approximately 50% of women had previously been screened for cervical cancer, and approximately 60% used a contraceptive, most commonly an injectable or implant. **Table 1**. Demographic and clinical characteristics of HPV+ Women Living with HIV assigned to Pocket or VIA study arm.

**Table 1.**
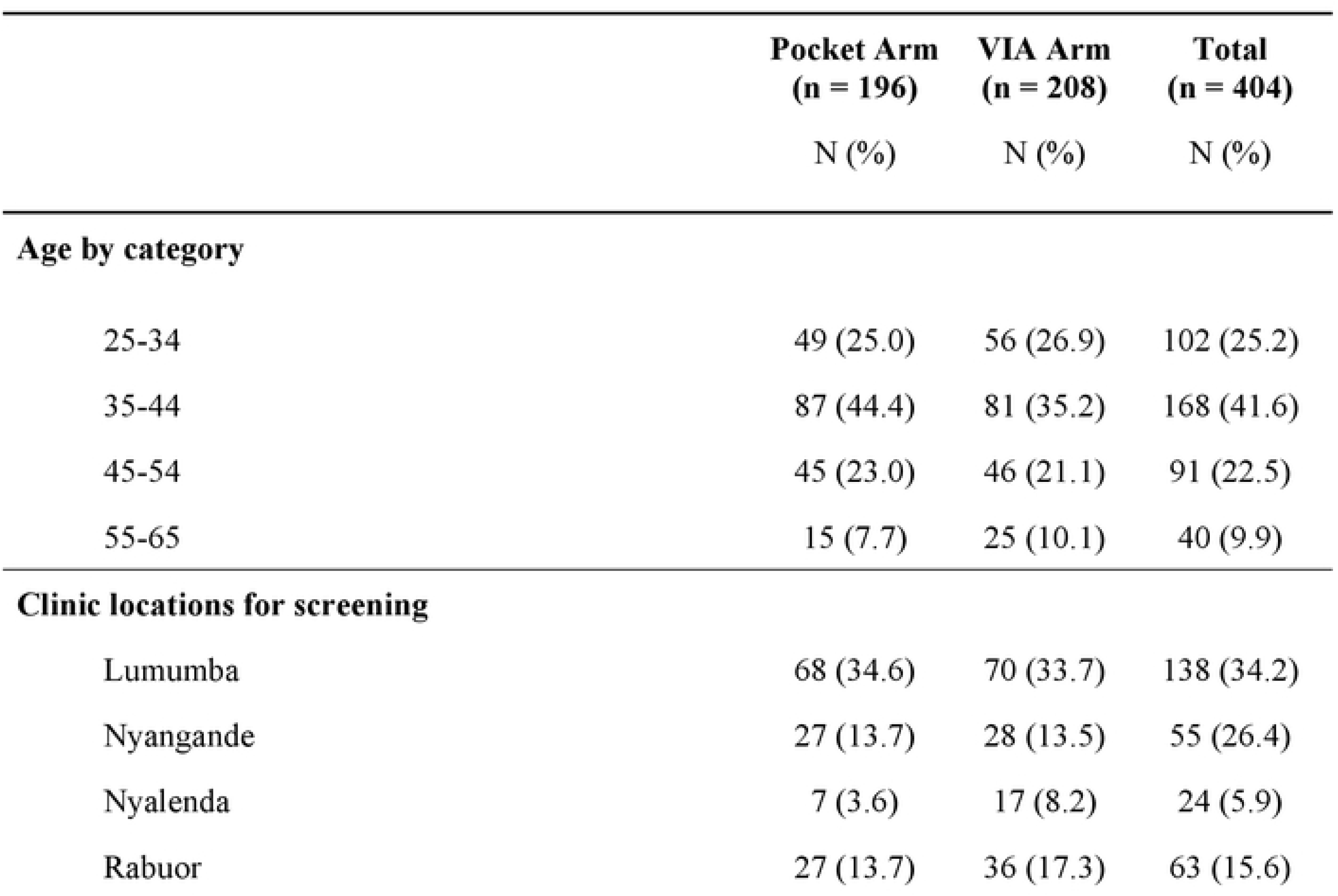
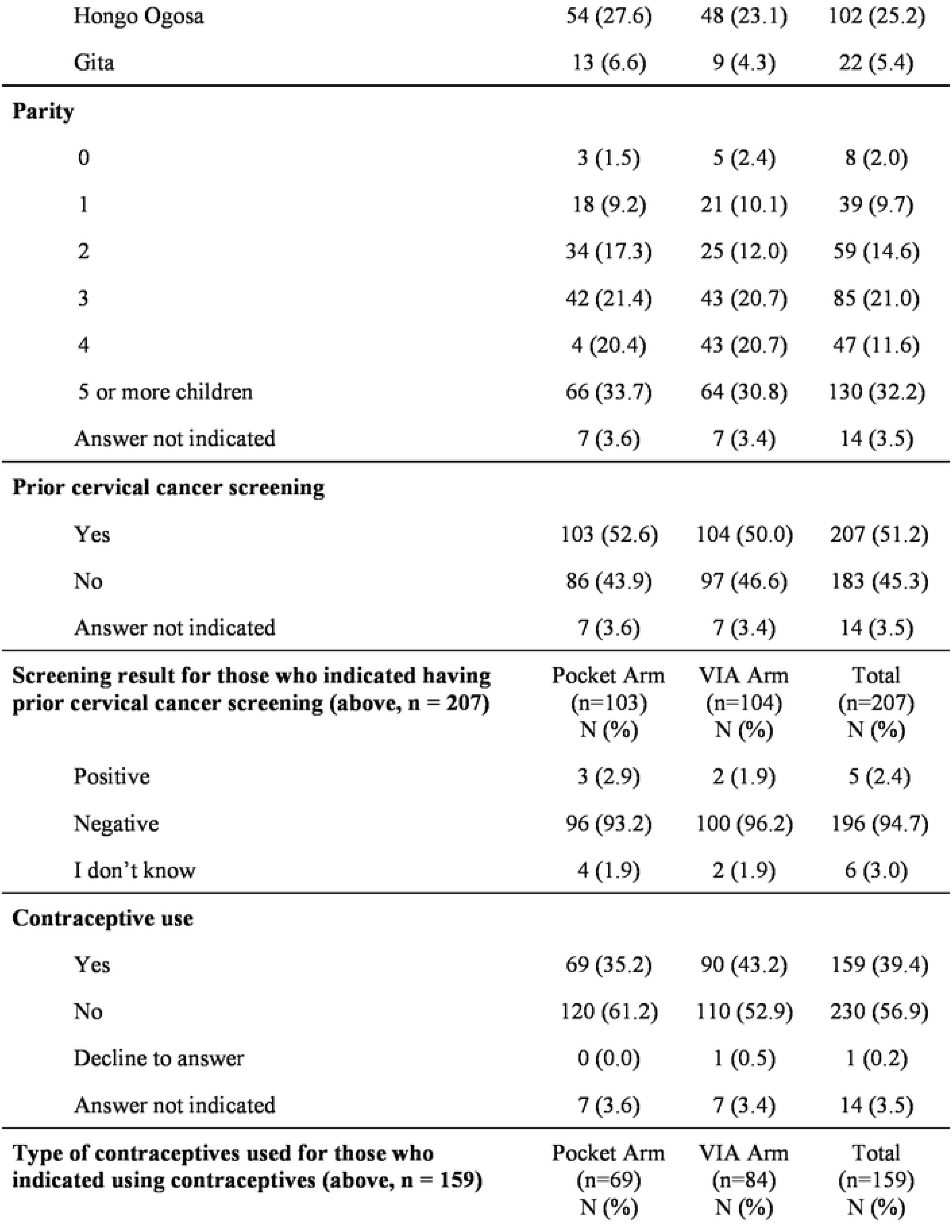
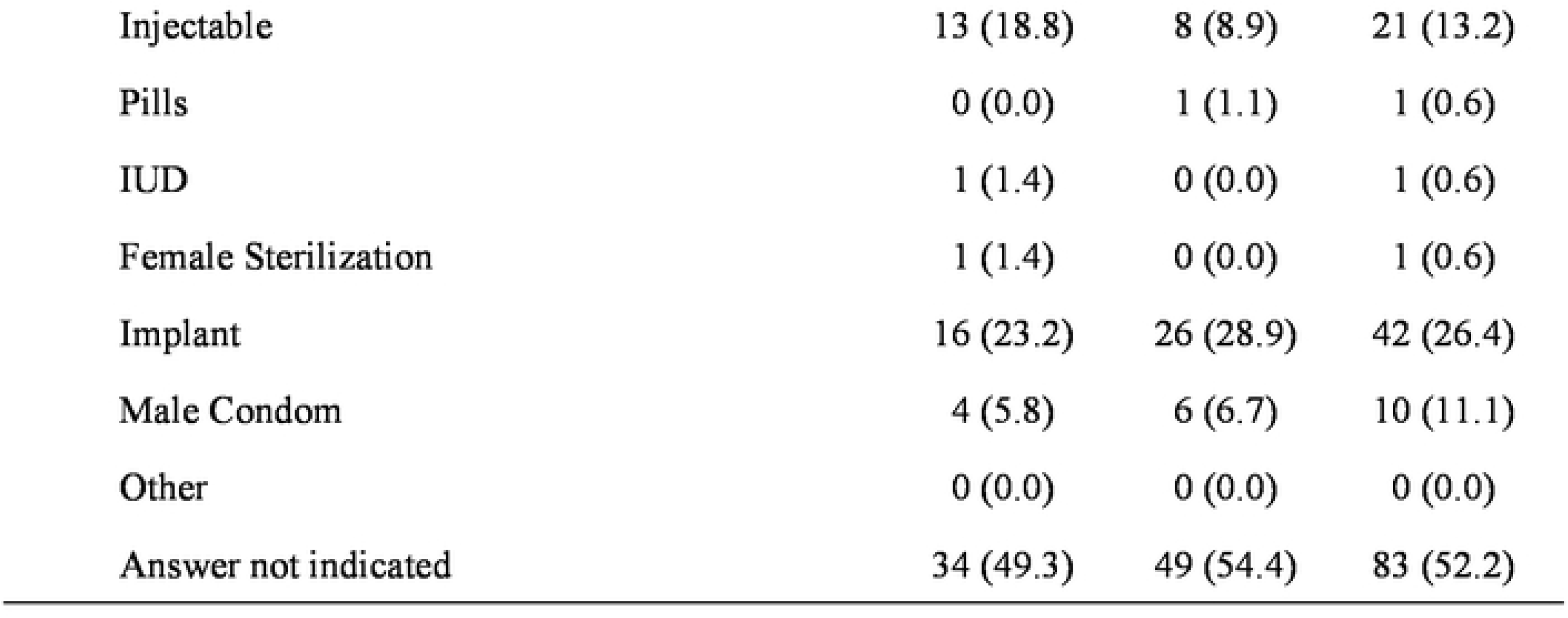

### Patient Diagnosis (Provider Visual Diagnosis and Histopathology)

In the Pocket arm, providers used the Pocket Colposcope to visualize the cervix before and after acetic acid application and to capture images using both white and green light (Figure 2). Only white light images were used to confer a diagnosis. The pre-acetic acid images as well as the green light images were captured for the purposes of algorithm development. It should be noted that at the time of data analysis, 388 out of 404 enrolled patients had processed biopsy results. The remaining 16 were lost during processing, resulting in a sample size of 196 and 197 for the Pocket arm and the VIA arm, respectively. The sample size in Tables 3,4, and 5 are consistent with that in Table 2. The percentage of a positive visual exam was 17.3% (95% CI = 12.4-23.1%) for the VIA arm compared to 14.3% (95% CI = 9.7-20.0%) for the Pocket arm (*X*^2^ = 0.48, p = 0.487; Figure 3). The overall rate of CIN2/3 confirmed on pathology was 15.5%, with 24 cases (12.2%) in the VIA arm and 36 cases (18.8%) in the Pocket Arm. There was a high rate of CIN3 for both the Pocket and VIA arms (17.3% and 9.6%, respectively) (Figure 3; Table 2). Majority of biopsies were histologically diagnosed as negative (NILM) in both the Pocket and VIA arms (75.9% and 80.2%, respectively). **Figure 3**. Representative cervix images of HPV+ women living with HIV, imaged with Pocket Colposcope at each stage of cervical precancer (pathology confirmed). **Table 2**. Provider and histopathology diagnoses following exams with Pocket Colposcope or VIA.

**Figure 3.**
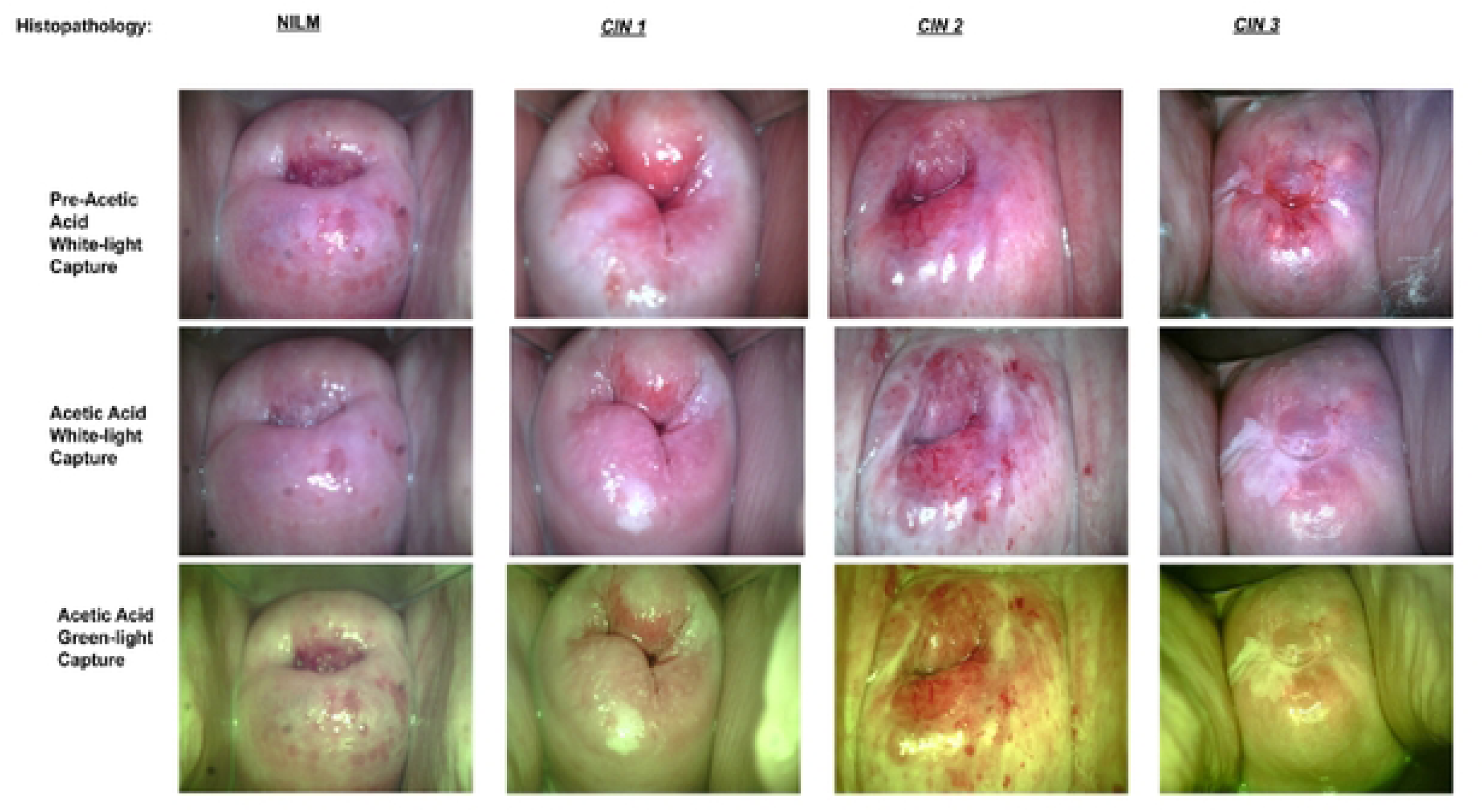

**Table 2.**

|  | Pocket Arm (n = 191) |  | VIA Arm (n = 197) |  | Total (n = 388) |
| --- | --- | --- | --- | --- | --- |
|  | N (%)<br>Total<br>patients | N (% of<br>pathology<br>category)<br>Correct<br>diagnosis | N (%)<br>Total<br>patients | N (% of<br>pathology<br>category)<br>Correct<br>diagnosis | N (%)<br>Total patients in<br>both arms |
| NILM | 145 (75.9) | 132 (91.0) | 155 (80.2) | 133 (85.8) | 300 (77.3) |
| CIN1 | 8 (4.2) | 4 (50.0) | 14 (7.1) | 9 (35.7) | 22 (5.7) |
| CIN2 | 3 (1.6) | 0 (0.0) | 5 (2.5) | 3 (60.0) | 8 (2.1) |
| CIN3 | 33 (17.3) | 10 (30.3) | 19 (9.6) | 3 (15.8) | 52 (13.4) |
| **CIN2/3** | **36 (18.8)** | **10 (27.8)** | **24 (12.2)** | **6 (25.0)** | **60 (15.5)** |
| SCC | 2 (1.0) | 0 (0.0) | 3 (1.5) | 1 (33.3) | 5 (1.3) |
| AGC | 0 (0.0) | N/A | 1 (0.5) | 0 (0.0) | 1 (0.3) |

*At the time of data analysis, 388 out of 404 enrolled patients had processed biopsy results. The remaining 16 were lost during processing resulting in a sample size of 191 and 197 for the Pocket arm and the VIA arm, respectively. The sample size in Tables 3, 4, and 5 are consistent with that in Table 2. NILM = negative for intraepithelial lesion or malignancy; CIN1, CIN2, CIN3 = cervical intraepithelial neoplasia grades 1-3; SCC = squamous cell carcinoma; AGC = atypical glandular cells*

The diagnostic characteristics were similar for VIA and Pocket with exception of the PPV. The sensitivity for correctly identifying CIN2+ (including CIN2, CIN3, SCC, and AGC) lesions was similar (26.3% vs. 25.0%, respectively; Table 3). The specificity of the Pocket arm was 4.9% higher than that of the VIA arm (88.9% vs. 84%, respectively; Table 3). The NPV for the VIA arm (87.1%) was approximately 4% higher than that of the Pocket arm (82.9%). On the other hand, the PPV for the Pocket arm (37%) was almost a factor of two higher than that of the VIA arm (20.6%). **Table 3**. Sensitivity, specificity, positive predictive value (PPV), and negative predictive value (NPV) for provider visual diagnosis of the cervix, compared to pathology.

**Table 3.**

|  | Pocket Arm (n = 191) | VIA Arm (n = 197) |
| --- | --- | --- |
| **Sensitivity** | 26.3% | 25.0% |
| **Specificity** | 88.9% | 84.0% |
| **PPV** | 37.0% | 20.6% |
| **NPV** | 82.9% | 87.1% |
| *Positive pathology diagnosis = HSIL/CIN2, HSIL/CIN3, SCC, AGC* |  |  |
| *Negative pathology diagnosis= NILM, LSIL/CIN1* |  |  |

### Provider and Participant Experience with Pocket Colposcope Compared to VIA

Study exams were carried out by 14 providers (Table 4). Each provider completed both Pocket and VIA exams based on which arm the patient was randomized to following consent. Majority of providers expressed “no challenges” during the exam for both procedures (96.0% and 97.1% for Pocket and VIA, respectively; Table 4). When using the Pocket, less than 1% of providers expressed difficulty with the internet, recording patient information/results, or perceived patient discomfort during a Pocket examination. Difficulty visualizing the patient’s cervix was expressed for 2.5% of Pocket exams compared to 0.5% of VIA exams. Additionally, providers answered that they were able to “complete the exam” and “make a diagnosis” for both the Pocket and VIA exams 100% of the time (Table 4).

**Table 4.**

|  | Pocket<br>(n = 191)<br>N (%) | VIA<br>(n = 197)<br>N (%) |
| --- | --- | --- |
| **8. Did any patient share their discomfort, concerns or pain during today's exam?** |  |  |
| Yes | 53 (27.7) | 53 (26.9) |
| No | 138 (72.2) | 144 (73.1) |
| **10. Did you run into any of the following challenges during today's exam?** |  |  |
| No challenges | 184 (96.3) | 191 (97.0) |
| Difficulty with internet | 1 (0.5) | N/A |
| Difficulty visualizing cervix | 5 (2.6) | 1 (0.5) |
| Difficulty recording patient info or results | 1 (0.5) | 0 (0) |
| Patient refusal | 0 (0) | 1 (0.5) |
| Patient discomfort | 1 (0.5) | 4 (2.0) |

A total of 190 women in the Pocket arm and 197 participants in the VIA arm completed the participant survey (Table 5). Almost all participants stated that the provider explained the importance of getting treated for cervical cancer and why they were either being examined by VIA or the Pocket (Table 5). Participants indicated little or no knowledge about cervical cancer and cervical cancer prevention prior to the exam, with majority expressing “none” or a “little” knowledge prior to the exam (Pocket: 94.8%, VIA: 95.9%). However, participants felt that their knowledge increased following the exam, with 81.0% of participants in the Pocket arm and 84.8% of participants in the VIA arm stating that they knew “somewhat” or “very much” about cervical cancer and its prevention.

**Table 5.**
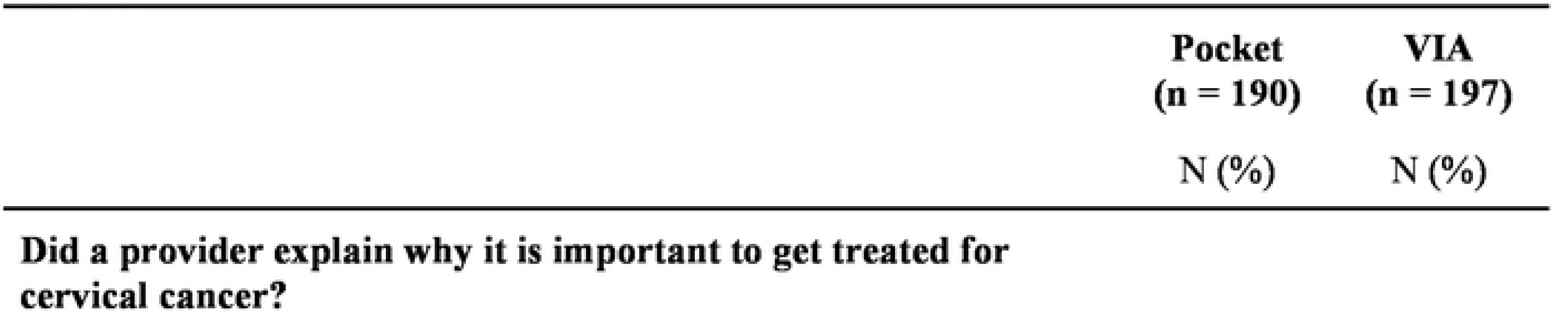
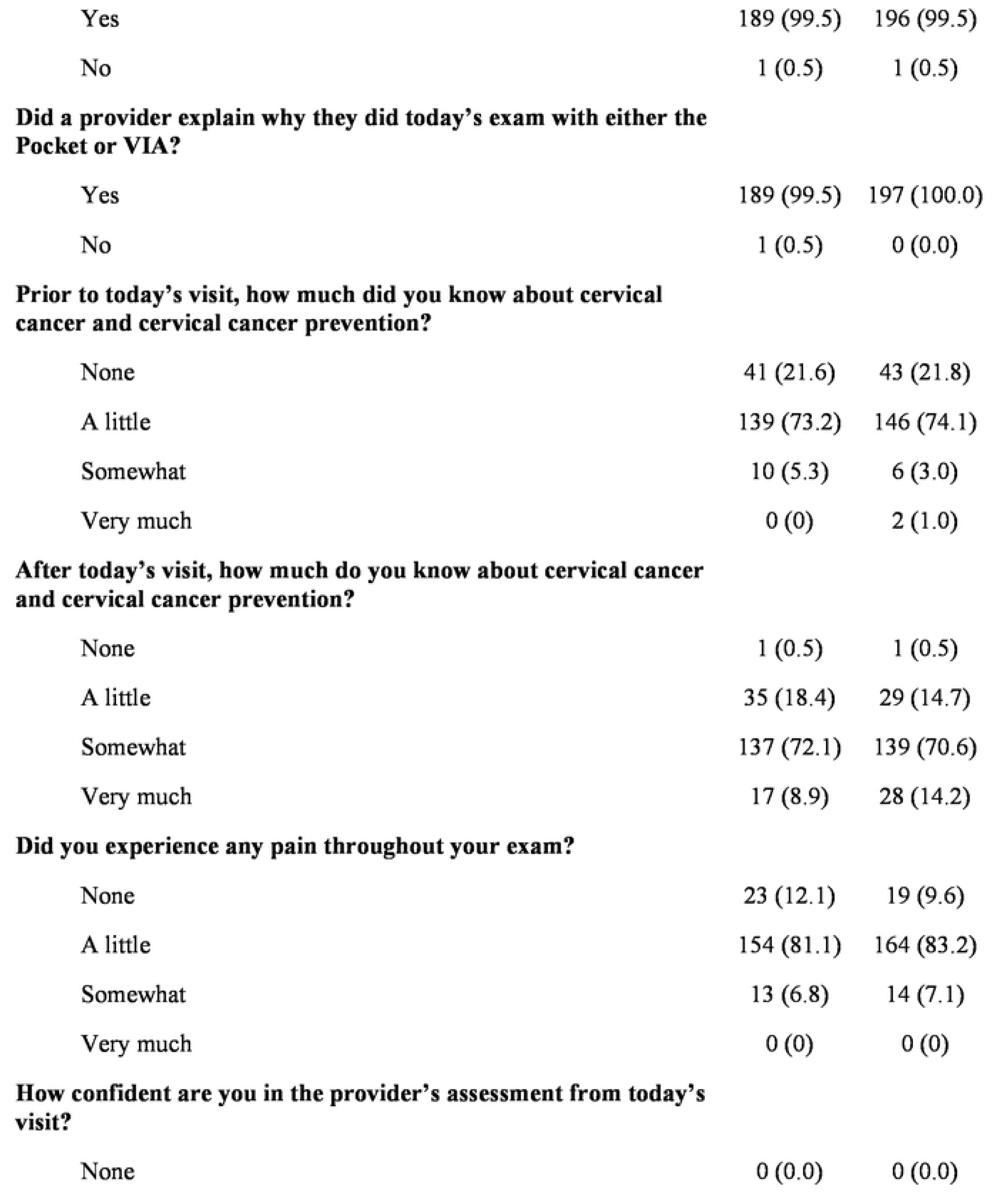
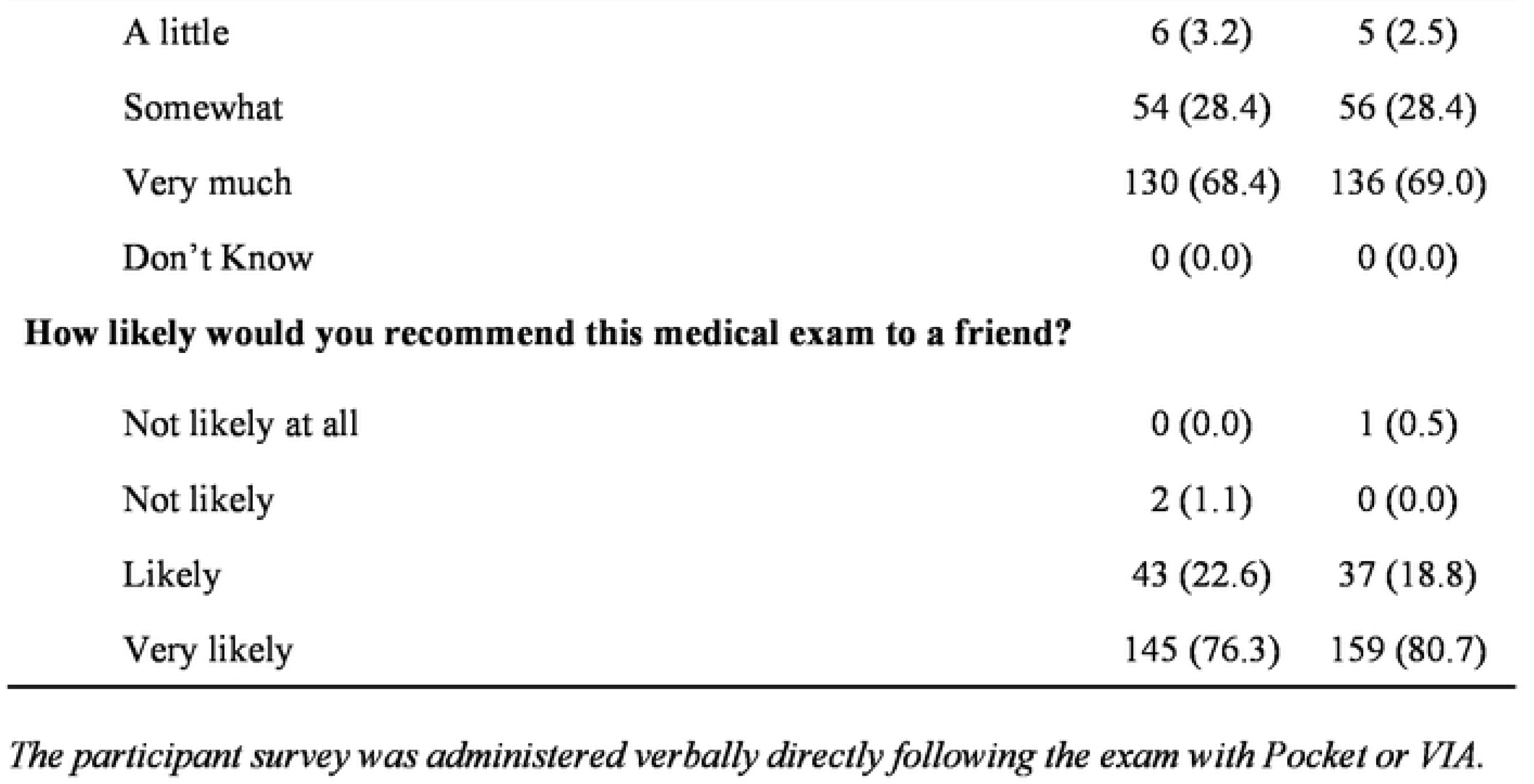

Providers answered that their patients shared their discomfort, concern, or pain during the examination with both the Pocket and VIA alone (27.7% and 26.9% respectively; Table 4). Consistently, majority of participants expressed little or no pain (Pocket: 93.2%, VIA: 92.8%; Table 5). Almost all participants in both arms stated that they were “somewhat” or “very much” confident about the providers’ assessment (Pocket: 96.8%, VIA: 97.4%) and similarly, most would “likely” or “very likely” recommend the exam to a friend (Pocket: 98.9%, VIA: 99.5%). **Table 4**: Post-exam Provider Pocket vs VIA Survey Question 8 and 10, full survey in Appendix. **Table 5**. Post-exam Female Participant Pocket vs. VIA Survey Responses, full survey in Appendix.

## Discussion

We sought to compare the diagnostic accuracy of VIA to the Pocket colposcope through an RCT of HPV positive women living with HIV (WLWH) in western Kenya. The results of this study show prevalence of CIN lesions among HIV-positive women consistent with other studies.(13)^-^(14) The study arm that used the Pocket colposcope had similar sensitivity, and a slightly lower specificity and NPV compared to that of VIA; however, it had almost a factor of two higher PPV. In this scenario, the Pocket Colposcope aided providers in yielding a greater percentage of pathology-confirmed positive visual diagnoses compared to VIA, without sacrificing sensitivity, thus avoiding unnecessary treatment. Providers expressed satisfaction with the use of the Pocket colposcope and reported very few challenges with the additional technology such as internet access and documenting information/results of the exam on the CHA app.

Majority of participants were satisfied with the exams in both arms, including with their provider’s explanation of the procedures and in the provider’s assessment, and would recommend the exam to a friend. They also expressed that their knowledge about cervical cancer screening and prevention increased after the exam. While very few participants reported pain during the survey, it is not surprising that a large percentage of women expressed some pain or discomfort during the exam, with no difference between arms. There are a number of studies that report that women fear the speculum-based exam and find that the speculum is painful and a deterrent for follow-up care.(15),(16),(17) The Pocket did not seem to add any additional discomfort or pain to the speculum exam.

VIA has been promoted as an effective strategy in low-resource settings because it is resource efficient and providers with limited colposcopy experience can be trained to complete the exam. One study in Kenya of WLWH showed that VIA had a sensitivity of 69.6% and specificity of 51% when CIN2+ lesions were used as the threshold.(14) Another study among WLWH in Kenya demonstrated VIA to have a sensitivity, specificity, PPV, and NPV of 86.6%, 71.6%, 30.3% and 97.4%, respectively for CIN2+ lesions as the threshold(18) Finally, one study conducted among WLWH in Nigeria reported a sensitivity, specificity, and PPV of VIA to be 76%, 83%, and 34% respectively for CIN2+ lesions as the threshold. The specificity of VIA in our study (84.0%) was comparable to or in some cases better than what has been reported previously; however, sensitivity (25.0%) was substantially lower. This is likely because none of these studies included HPV screening. On the other hand, the PPV (20.6%) and NPV (87.1%) in our study were more comparable with that reported previously.(19)

It is important to highlight that in general, direct comparison between different studies and ours is difficult owing to different populations, study designs, device characteristics, and provider training. Therefore, comparison between different approaches within the same study can potentially reduce these inconsistencies. Several investigators have compared VIA based on the unaided eye and imaging, as we have reported in this study. It is important to note that none of these studies were carried out on a HIV+ population. Comparison between VIA and the Cervioscope without HPV screening (a 13.6 MP Sony cyber shot camera) resulted in a sensitivity, specificity, PPV and NPV of 71.42%, 92%, 20%, and 99.1%, for VIA and 71.42%, 93.62%, 23.8%, and 99.1% for Cervioscope, when compared to histopathology.(20) Another study that compared VIA to digital colposcopy had different findings. The sensitivity, specificity, PPV, and NPV were 62.5%, 98.8%, 90.9%, and 93.2% for VIA and 46.7%, 97.6%, 77.8%, 91%, for digital colposcopy, and similar to the case in the previous study, no HPV test was performed as a primary screening step.(21) Surprisingly, both sensitivity and specificity were lower for the imaging arm. In another study, one gynecologist in Madagascar was asked to complete VIA for HPV+ women and then smartphone images were sent to 3 expert colposcopists in Geneva for classification. Interestingly, the comparison of groups did not reach significance due to small sample size, but the results were suggestive of the potential of digital colposcopy.(22) It is expected that any two-step screening process, such as HPV followed by VIA or Pocket, will have a lower sensitivity than a single step process since we are able to more effectively reduce false positives.

The results from our RCT study showed consistently lower sensitivity of 25-30% in both the VIA and Pocket colposcope arms. Much of this decrement in sensitivity is the fact that the visual exams were performed as a triage step after an initial HPV test, compared to the cited studies in which the visual assessment was the primary screening exam. However, this study has provided much information about the need for improved provider training and continued feedback and mentorship on exam and diagnostic quality. Because we sought to test the Pocket in a real-world clinical setting in Kenya, we engaged providers with varying levels of colposcopic expertise and experienced a high rate of provider turnover unrelated to the study. This is further supported by the fact that in our group’s previous studies, in which we found similar sensitivities and specificities between expert interpretation (by the same providers) of the Pocket colposcope and standard-of-care colposcope images (sensitivity and specificity was 79.8% and 56.6%, respectively for the standard of care colposcope and 71.2% and 57.5%, respectively for Pocket colposcope images).(12)

Ultimately, our study and those reported by others point to the importance of standardizing diagnostic interpretation, and there is a concerted effort in augmenting visual interpretation with machine learning algorithms. However, the ability to capture non-blurry images of all four quadrants of the cervix with minimal glare is equally important as any algorithm will be influenced by these factors.(23) A different study analyzing a portion of the data set used in the present study used an automated blur detection algorithm to assign binary categories of “Blurry” or “Clear” to each Pocket image.(24) This study found that an expert provider’s confidence in identifying a lesion was significantly higher in “Clear” versus “Blurry” images.(25) Another study assessing an automated diagnostic algorithm for cervix images taken with a mobile phone camera found that de-blurring images improved algorithm performance by 21.4%.(26) In addition to blur, other confounding factors, such as glare, can impact diagnostic interpretation. Robust, standardized, and continuous training on image interpretation and image quality will continue to be an important factor in the hands of less experienced providers. Automated methods/algorithms that provide real time or near real time feedback on the quality of the image during the exam can allow the provider to retake images, potentially improving quality control.

Overall, we found the Pocket Colposcope to be acceptable to providers and patients for clinic-based triage of HPV positivity in western Kenya. We did not find that Pocket colposcope substantially increased specificity or sensitivity over VIA when used as a visual adjunct alone though PPV was significantly higher reducing overtreatment. However, this study does provide key data to support use of Pocket as a feasible, lower cost colposcopic device, which could facilitate biopsy-confirmation of disease. The Pocket can also be explored as a mode of increased provider training, patient education or a way to facilitate remote diagnosis. Finally, the images and image quality feedback can be used to inform algorithms for image improvement or automated algorithms for diagnosis of precancer.

## Data Availability

All data is available within Duke’s research data repository.

## Acknowledgments

The team would like to acknowledge the significant efforts of the Kenya Medical Research Institute (KEMRI). Jeniffer Ambaka, study coordinator, Saduma Ibrahim, Data Manager, Evans Obuto and the research assistants.

